# Cost-Effectiveness of Prefusion F Protein-Based Vaccines Against Respiratory Syncytial Virus Disease for Older Adults in the United States

**DOI:** 10.1101/2023.08.14.23294076

**Authors:** Seyed M. Moghadas, Affan Shoukat, Carolyn E. Bawden, Joanne M. Langley, Burton H Singer, Meagan C. Fitzpatrick, Alison P. Galvani

## Abstract

**Background:** Two prefusion F protein-based vaccines, Arexvy and Abrysvo, have been authorized by the US Food and Drug Administration for protecting older adults against Respiratory Syncytial Virus (RSV)-associated lower respiratory tract illness. We evaluated the health benefits and cost-effectiveness of these vaccines.

**Methods:** We developed a discrete-event simulation model, parameterized with the burden of RSV disease including outpatient care, hospitalization, and death for adults aged 60 years or older in the US. Taking into account the costs associated with these RSV-related outcomes, we calculated the net monetary benefit using quality-adjusted life-years (QALY) gained as a measure of effectiveness, and determined the range of price-per-dose (PPD) for Arexvy and Abrysvo vaccination programs to be cost-effective from a societal perspective.

**Results:** Using a willingness-to-pay of $95,000 per QALY gained, we found that vaccination programs could be cost-effective for a PPD under $120 with Arexvy and $111 with Abrysvo over the first RSV season. Achieving an influenza-like vaccination coverage of 66% for the population of older adults in the US, the budget impact of these programs at the maximum PPD ranged from $5.74 to $6.10 billion. If the benefits of vaccination extend to a second RSV season as reported in clinical trials, we estimated a maximum PPD of $250 for Arexvy and $233 for Abrysvo, with two-year budget impacts of $11.59 and $10.89 billion, respectively.

**Conclusions:** Vaccination of older adults would provide substantial direct health benefits by reducing outcomes associated with RSV-related illness in this population.

## Introduction

Respiratory syncytial virus (RSV) is a major cause of lower respiratory tract disease (LRTD) among older adults [1–3], with significant health and socio-economic burden on a global scale [4]. RSV-related hospitalizations and healthcare costs are exacerbated by the presence of comorbidities, the rates of which have been increasing among older adults [1]. In the United States (US) alone, the annual direct and indirect costs of RSV disease in adults aged 60 years or older are estimated to exceed $3.9 billion [5]. To reduce the burden of RSV disease among older adults, two highly efficacious prefusion F protein vaccines (Arexvy and Abrysvo) [6,7] have been developed, authorized by the US Food and Drug Administration, and recommended by the Centers for Disease Control and Prevention [8,9].

With the availability of these vaccines, determining vaccination strategies that are cost-effective remains an important component of program implementation. In this study, we conducted a cost-effectiveness analysis of various RSV vaccination programs by developing a discrete-event probabilistic model of RSV outcomes for adults aged 60 years and older in the US. The model includes important characteristics of the study population with estimates of disease burden and the associated costs. Using stochastic simulations, we calculated the net-monetary benefit (NMB), the incremental cost-effectiveness ratio (ICER), and the budget impact associated with programs evaluated. In addition, we determined the range of vaccine price-per-dose (PPD) within which a program would be cost-effective, while accounting for the reported efficacy estimates of Arexvy and Abrysvo against RSV LRTD. Considering direct and indirect costs of RSV disease outcomes and management, we performed our analysis from a societal perspective.

## Methods

### Model structure and study population

We developed a discrete-event simulation model (**Figure 1**) with a population of 100,000 adults stratified into age groups of 60 to 64, 65 to 69, 70 to 74, 75 to 79, 80 to 84, and 85 years or older reflecting US demographics [10]. We considered the prevalence of comorbidities across these age groups (**Supplementary Table A1**) [11], which was used in determining the severe outcomes of RSV disease for adults with 0, 1–3, and ≥4 comorbidities [12].

**Figure 1.**
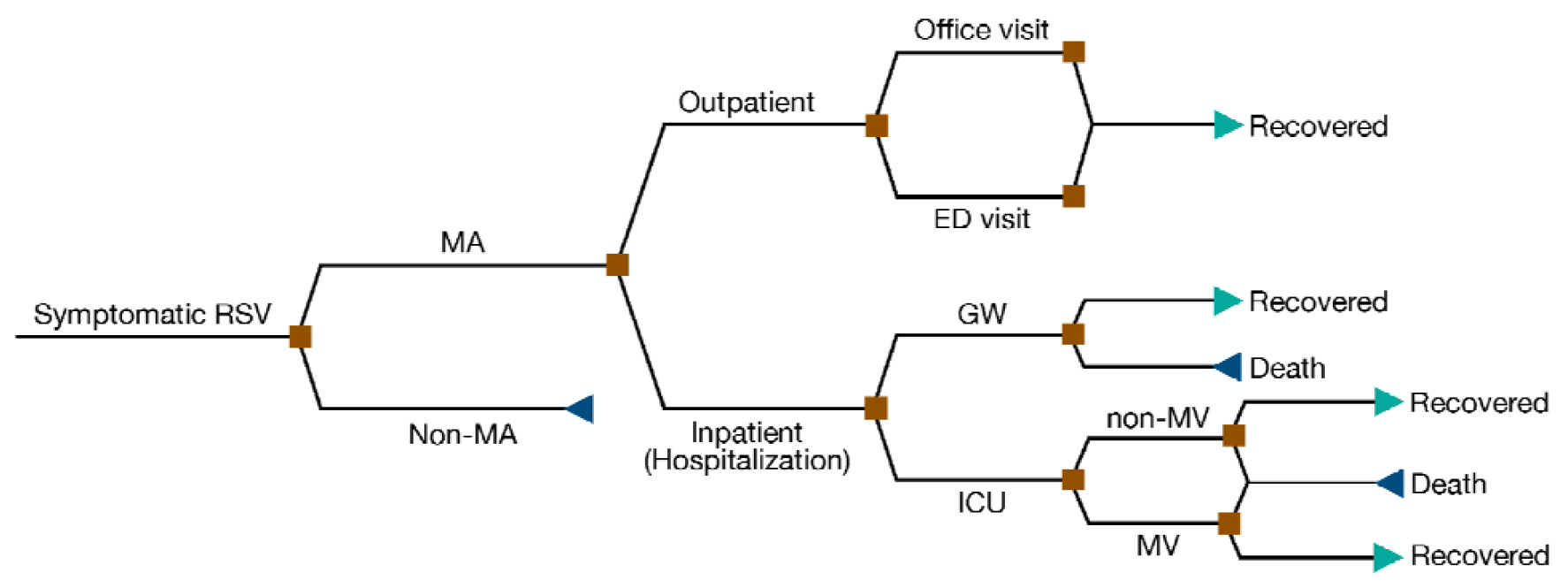
Structure of the discrete-event simulation model applied to scenarios in the absence and presence of interventions for different outcomes. MA: medically attended; ED: emergency department; GW: general ward; ICU: intensive care unit; MV: mechanical ventilation.

### RSV-related outcomes

The model was parameterized with sampled annual incidence of medically-attended (MA) RSV cases per 100,000 adults aged 60 years or older, stratified as outpatient (i.e., office visits and emergency department (ED) visits) and inpatient (i.e., hospitalization in the general ward, and intensive care unit (ICU) admissions) [13]. We unformly sampled the annual incidence of outpatient office visits from the range 1595–2669, with a mean of 2133. Annual ED visits were uniformly sampled from the range 23–387, with a mean of 201. The annual incidence of hospitalizations, including ICU admissions, was sampled uniformly from the range 178–250, with a mean of 214. Distribution of hospitalizations among age groups were parameterized based on the average rates reported by RSV-NET for four seasons from 2016-17 to 2019-20 [14]. The proportions of hospitalized cases were 6.2%, 12.6%, 26.5%, and 54.8% among age groups 60–64, 65–74, 75–84, and 85 years or older, respectively. Among hospitalized patients, 5.5% had no comorbid conditions, 78.2% were with 1–3 comorbidities, and 16.3% had ≥4 comorbidities [12]. The rates of ICU admission were set to 24%, 15%, and 12% for patients with 0, 1–3, and ≥4 comorbidities, respectively [12]. Among patients admitted to the ICU, 16.6% required the use of mechanical ventilation (MV) [14]. The death rate for hospitalized patients was sampled uniformly in the range 5.6%–7.6% [15], distributed asf 37% from the general ward and 63% from those admitted to ICU [12].

The duration of symptomatic RSV disease for outpatient care was sampled uniformly between 7 and 14 days [15]. For RSV-related hospitalization, we included a 4-day delay between the onset of symptoms and hospital admission [16,17]. The duration of hospitalization in the general ward was sampled from a Gamma distribution, with a mean of 6.2 days and standard deviation of 5.6 days [18]. For patients admitted to ICU, we assumed a 1-day interval between hospital admission and ICU admission [19]. The length of ICU stay was sampled from a Gamma distribution with a mean of 4.5 days and standard deviation of 3.6 days [20]. For hospitalized patients who survived, we assumed 2 days in the general ward post-ICU [21].

### Costs of RSV-related outcomes

Direct costs of RSV-related outcomes included office visits, emergency department (ED) visits, and hospitalization (**Table 1**). For indirect costs, productivity loss was calculated for the duration of RSV disease and associated outcomes, as well as the lost income for the remaining life expectancy for those patients who died (**Supplementary Table2 A1, A2**). For the short-term loss of productivity, we considered the age-stratified proportion of the study population participating in the labor force (**Supplementary Table A2**) [22], and used the median income of $63,336 for the 60-64 years age group, and $54,184 for working adults aged 65 years or older for 2023 (**Supplementary Table A2**) [23]. These estimates correspond to daily wages of $151.6, $52.7, and $18.1 for 22 working days per month among working adults in the age groups 60–64, 65–74, and 75 years or older, respectively. To calculate the total monetary loss in the event of death due to RSV, we used estimates of remaining life for different age groups (**Supplementary Table A1**) [24], with an annual discounting rate of 3%. All costs were converted and inflated to 2023 US dollars.

**Table 1.** Model parameters for the duration of RSV-related outcomes and associated costs.

| RSV-related outcome | Duration | unit | Source |
| --- | --- | --- | --- |
| Symptomatic disease, non-MA<sup>a</sup> RSV cases | 2–8 | days | [32] |
| Symptomatic disease, MA RSV outpatient care | 7–14 | days | [15] |
| Median time interval between onset of symptoms and hospital admission | 4 | days | [16,17] |
| Length of hospitalization in GW<sup>b</sup> | Mean: 6.2<br>Gamma(1.2258, 5.0582) | days | [18] |
| Length of hospitalization in ICU | Mean: 4.5<br>Gamma(1.5625, 2.8802) | days | [20] |
| RSV-related outcome | Cost estimates | Unit | Source |
| Office visit | \$126 | Per visit | [15] |
| ED visit | \$649 | Per visit | [12] |
| Hospitalization, GW | \$1,116 | Per diem | [12,15,33,34] |
| ICU without MV<sup>c</sup> | \$3,348 | Per diem | |
| ICU with MV | \$4,870 | Per diem | |

<sup>a</sup> MA: medically attended; <sup>b</sup> GW: general ward; <sup>c</sup> MV: mechanical ventilation

### RSV vaccination strategies and associated costs

We considered two scenarios of RSV vaccination coverage of older adults. In the first scenario (S1), we assumed a coverage of 66% similar to the average influenza vaccination coverage from 2010-11 to 2020-21 seasons in the US for adults aged 65 years or older [25]. For comparison purposes, we also considered a second scenario (S2) with a 100% vaccination coverage for this population. Based on the seasonality of RSV in the US (**Supplementary Figure A1**) [26], we assumed vaccination beginning in September (similar to timelines for influenza seasonal vaccination), achieving the target coverage within 2 months in each scenario.

For this analysis, we varied the purchasing cost of a single dose of RSV Prefusion F Protein vaccine between $50 and $500 to determine the price range within which an immunization program would be cost-effective. The average cost for administering a vaccine was set to $25 per dose [27].

### Efficacy of RSV vaccines

We considered two RSV prefusion F protein vaccines authorized in the US, Arexvy from GlaxoSmithKline and Abrysvo from Pfizer. The efficacy of a single dose of Arexvy against MA RSV-related LRTD, applied as efficacy against outpatient care in this study, was estimated at 82.6% (95% confidence interval [CI], 57.9% — 94.1%) over a median follow-up period of 6.7 months [6]. Arexvy efficacy against severe RSV-related LRTD, applied as efficacy against hospitalization in this study, is estimated at 94.1% (95% CI, 62.4% — 99.9%) [6]. Similarly, the efficacy of Abrysvo against MA RSV-related LRTD, applied as efficacy against outpatient care in this study, is estimated at 65.1% (95.0%% CI: 35.9% — 82.0%) through the end of the first RSV season [7]. Abrysvo efficacy against severe RSV-related LRTD, applied as efficacy against hospitalization in this study, is estimated at 88.9% (95.0% CI: 53.6% — 98.7%) [7].

For cost-effectiveness analysis of these vaccines and to account for waning immunity, we considered two profiles of temporal decay for vaccine efficacy corresponding to sigmoidal and constant. For the first profile, we fitted a sigmoidal function over a 24-month period to derive point estimates with the same mean efficacy as estimated in clinical trials (**Supplementary Figure A2**). For Arexvy, we considered a sigmoidal decay over 24 months post vaccination and used an 18-month follow up period with estimates of 67.2% (95% CI: 48.2% — 80.0%) against outpatient care and 78.8% (95% CI: 52.6% — 92.0%) against hospitalization, derived in the secondary endpoint analysis [6]. Similarly, for Abrysvo we fitted a sigmoidal decay of immune protection across 24 months post vaccination, and used secondary estimates of 48.9% (95.0% CI: 13.7% — 70.5%) against outpatient care and 78.6% (95.0% CI: 23.2% — 96.1%) against hospitalization during 18 months follow-up [7]. For the constant profile, we used efficacy estimates as reported in clinical trials over the follow-up periods, with a linear decline beginning at 18 months post vaccination (**Supplementary Figure A3**).

### Cost-effectiveness analysis

To conduct a cost-effectiveness analysis, we calculated the net monetary benefit as NMB = Δ*E* × WTP − Δ*C*, where Δ*E* represents the quality-adjusted life-year (QALY) saved using vaccination compared to no intervention, Δ*C* is the incremental costs associated with the vaccination scenario, and WTP is the willingness-to-pay threshold for a QALY gain. A vaccination scenario was considered cost-effective if it resulted in a positive NMB. In our analysis, we calculated the monetary value of health gained using a WTP threshold of $95,000 per QALY [28]. We also estimated the ICER for each vaccination scenario as Δ*C/*Δ*E* to measure the additional costs incurred for gaining one QALY. Total QALYs in each scenario were calculated based on the health utility values related to RSV disease and outcomes among different age groups in the study population (**Supplementary Table A3**) [15,24,29]. Cost-effectiveness analyses were conducted from a societal perspective considering both direct and indirect costs.

For the primary analysis, we considered a time horizon of a single (first) RSV season. In the secondary analysis, the time horizon was set to two RSV seasons in light of the efficacy estimates for a single dose over a 24-month period (**Supplementary Data**).

### Model Implementation

The model was simulated stochastically using Monte-Carlo sampling for a total of 1000 independent realizations. In each realization, model parameters (**Table 1**) were sampled for each individual from their respective distributions or estimated ranges, thus probabilistically accounting for the sensitivity of the outcomes with respect to input values. To generate 95% confidence intervals around point estimates, we employed a nonparametric, bias-corrected and accelerated bootstrap technique with 1000 replicates. The cost-effectiveness analysis was conducted for both a single season and two seasons, comparing vaccination scenarios S1 and S2 to the scenario with no intervention. For each scenario of vaccination, we considered three cases for the use of vaccines: (i) Arexvy alone, (ii) Abrysvo alone, and (iii) a combination of Arexvy and Abrysvo with a probability of 50% receiving one of these vaccines to achieve the target coverage. For case (iii), we assumed that the PPD would be the same for both vaccines. The computational model is available at: https://github.com/affans/rsv-ce-adults.

### Ethics and guidelines

This study used published estimates and publicly available data sources and thus no ethics approval was required. Consolidated Health Economic Evaluation Reporting Standards (CHEERS) for reporting were followed [30].

## Results

### Health outcomes

Using the sigmoidal vaccine efficacy profile (**Supplementary Figure A2**), Scenario S1 with 66% vaccination coverage resulted in a mean reduction of 53.6%, 41.5%, and 47.6% in outpatient care using Arexvy only, Abrysvo only, and a combination of Arexvy and Abrysvo, respectively, for the first RSV season (**Figure 2A**). The corresponding reductions in hospitalizations were 60.0%, 57.7%, and 59.1%. RSV-related deaths were reduced by 60.5%, 58.4%, and 58.9%. Increasing vaccination coverage to 100%, S2 resulted in mean reductions of 81.2%, 63.0%, and 72.1% in outpatient care; 91.6%, 87.4%, and 89.5% in hospitalizations; and 91.3%, 88.1%, and 89.7% in deaths using Arexvy only, Abrysvo only, and a combination of Arexvy and Abrysvo, respectively (**Figure 2B**). When constant vaccine efficacy profiles were used (**Supplementary Figure A3**), we found an insignificant change in the reduction of outcomes compared to the sigmoidal vaccine efficacy profiles over the first RSV season (**Figure 2C-D**). The age-specific reduction of outcomes were similar to the overall reduction in the corresponding scenarios of S1 and S2 (**Supplementary Figures A4, A5**).

**Figure 2.**
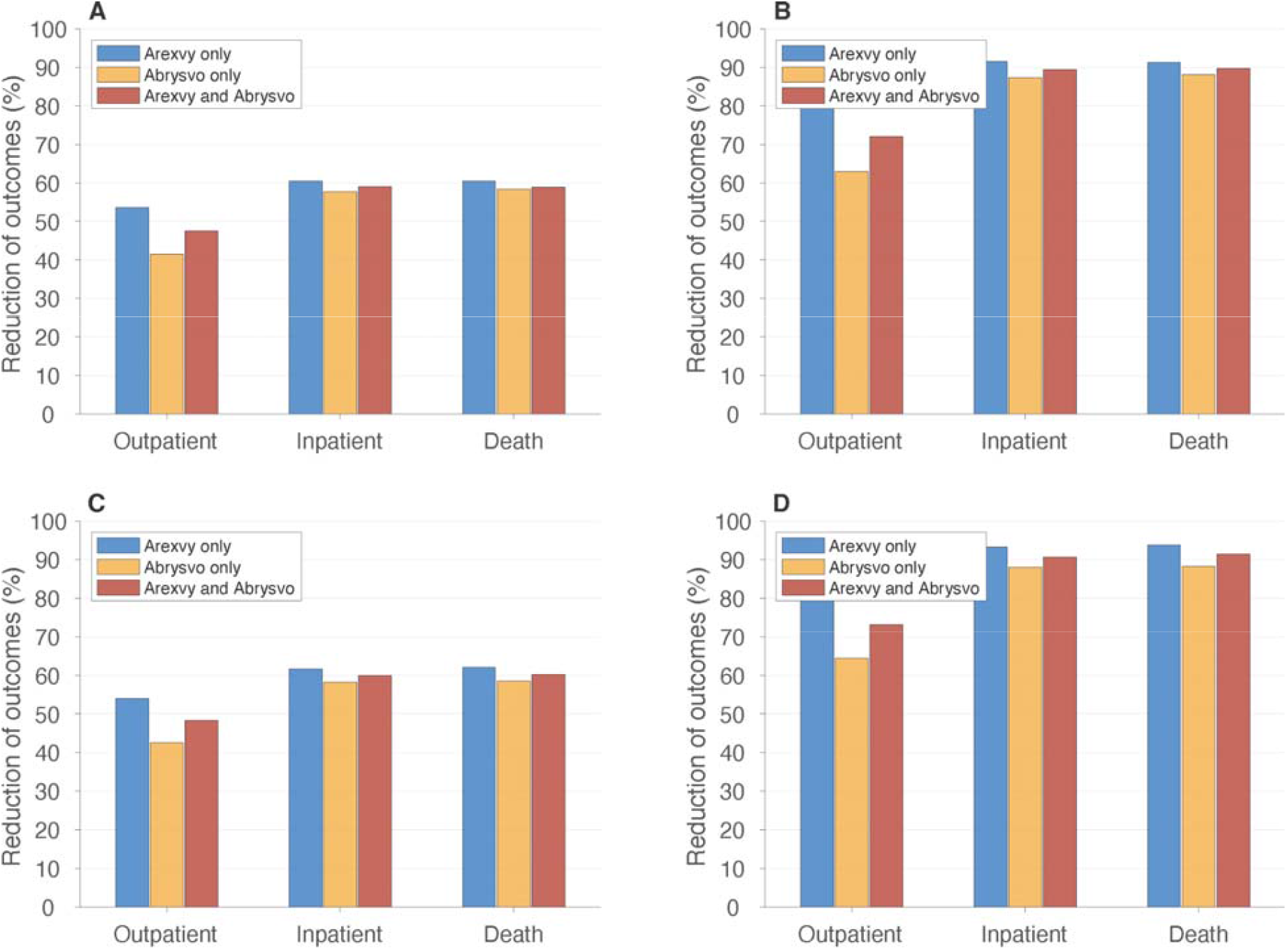
Overall reduction of RSV-related outpatient care (office and ED visits), inpatient care (hospitalization), and death among adults 60 years of age or older, compared to the scenario without vaccination, with sigmoidal (A,B) and constant (C,D) vaccine efficacy profiles. Vaccination coverage was set to 66% (A,C) and 100% (B,D).

### Number needed to vaccinate (NNV)

The mean number of adults in the study population who needed to be vaccinated to avert one outpatient visit during the first RSV season with sigmoidal vaccine efficacy profiles ranged from 53 to 60 (**Supplementary Table A4**). Mean estimated NNV to avert one hospitalization ranged from 511 to 538, and from 7,788 to 8,108 to prevent one death. Similar ranges of NNV were found using constant vaccine efficacy profiles (**Supplementary Table A4**). At the national level with a population of approximately 74 million older adults in the US, these findings indicate that 66% vaccination coverage of adults aged 60 years or older could avert over 810,000 outpatient (office and emergency department) visits, 90,500 hospitalizations, and 6,000 deaths associated with RSV disease over the first RSV season.

### Cost-effectiveness of vaccination scenarios

We determined the maximum PPD below which programs with Arexvy, Abrysvo, or combination of both vaccines would be cost-effective (i.e., NMB>0) at a WTP of $95,000 per QALY gained over the course of the first RSV season (**Table 2**). Using sigmoidal vaccine efficacy profiles (**Supplementary Figure A2**), the maximum PPD for a positive NMB was $120 for Arexvy only, $111 for Abrysvo only, and $114 for combination of both vaccines at 66% vaccination coverage (**Figure 3A**). With 100% vaccination coverage, the corresponding maximum PPDs were $119, $110, and $116.

**Table 2.**
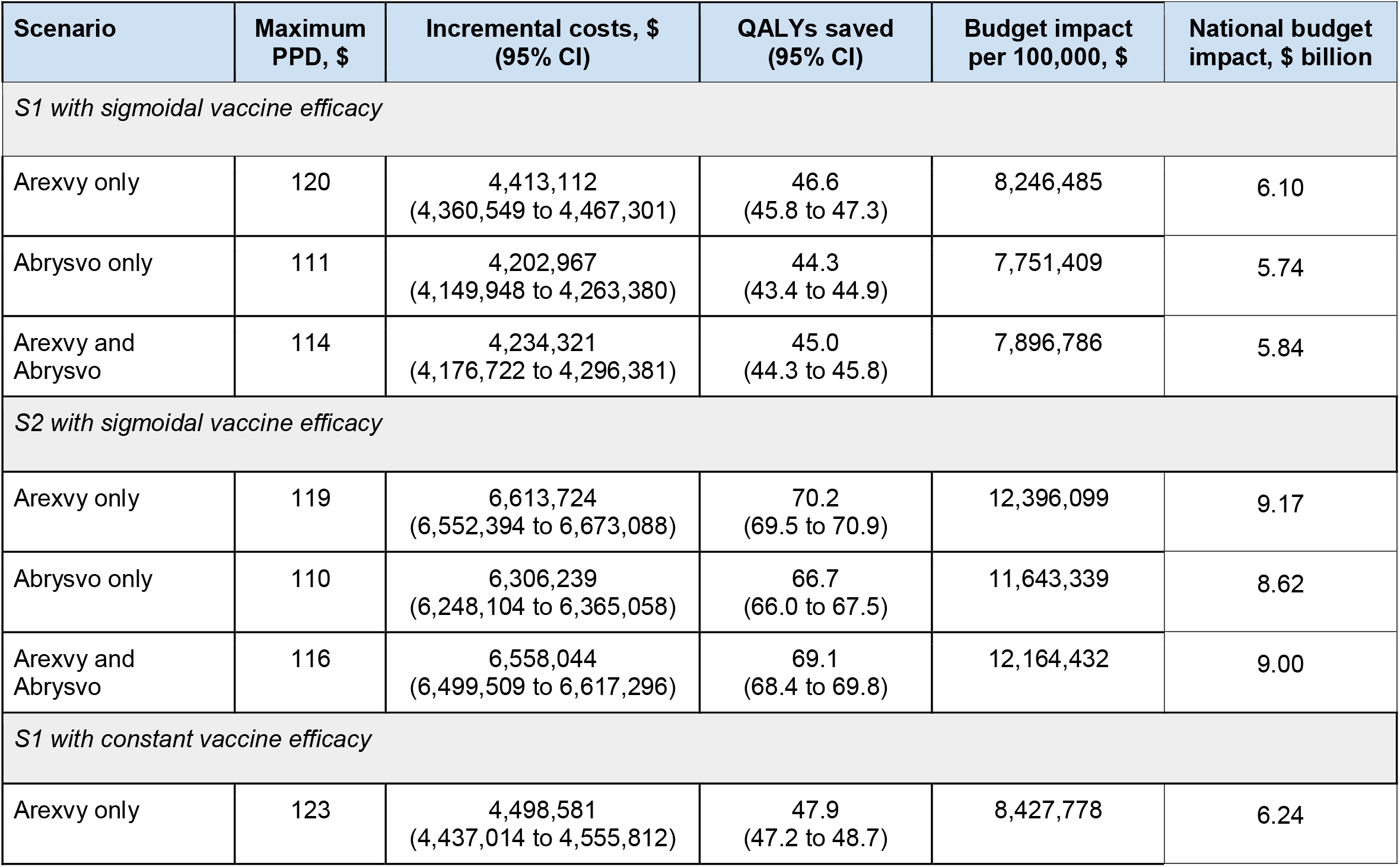
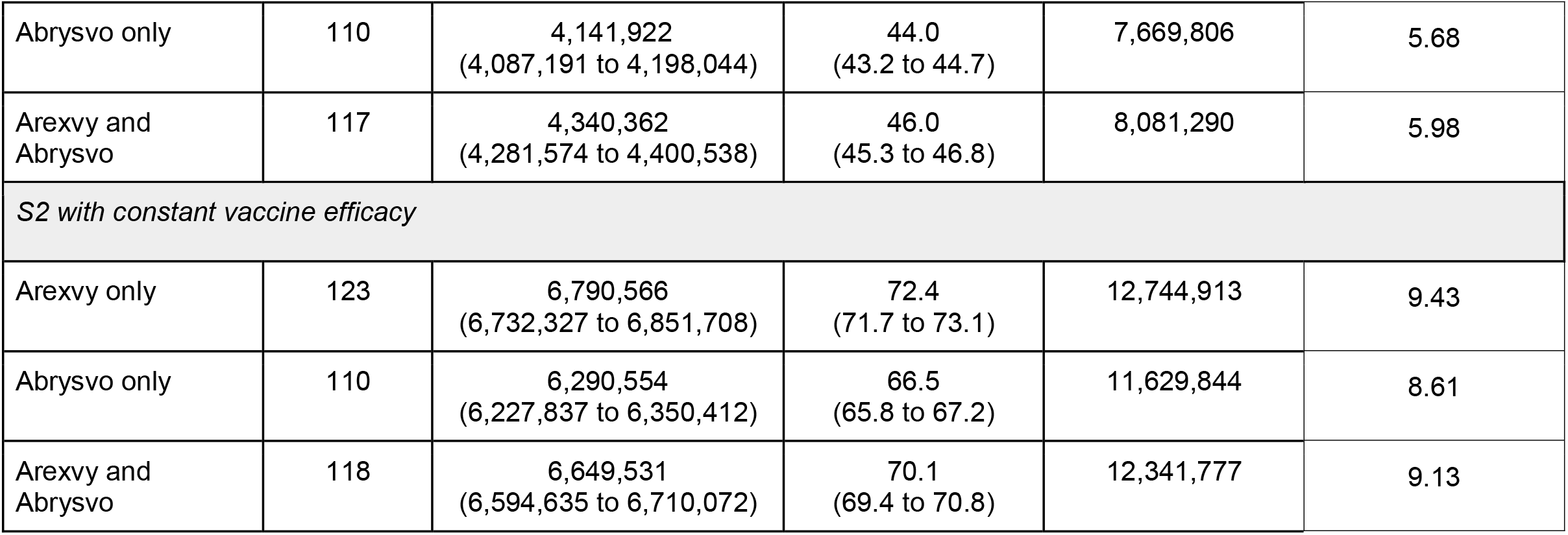
Model estimates of cost-effectiveness analyses for vaccination programs with Arexvy only, Abrysvo only, and combination of Arexvy and Abrysvo over the first RSV season in a population of 100,000 adults aged 60 years or older at the WTP of $95,000. All strategies were compared to the baseline with no intervention.

**Figure 3.**
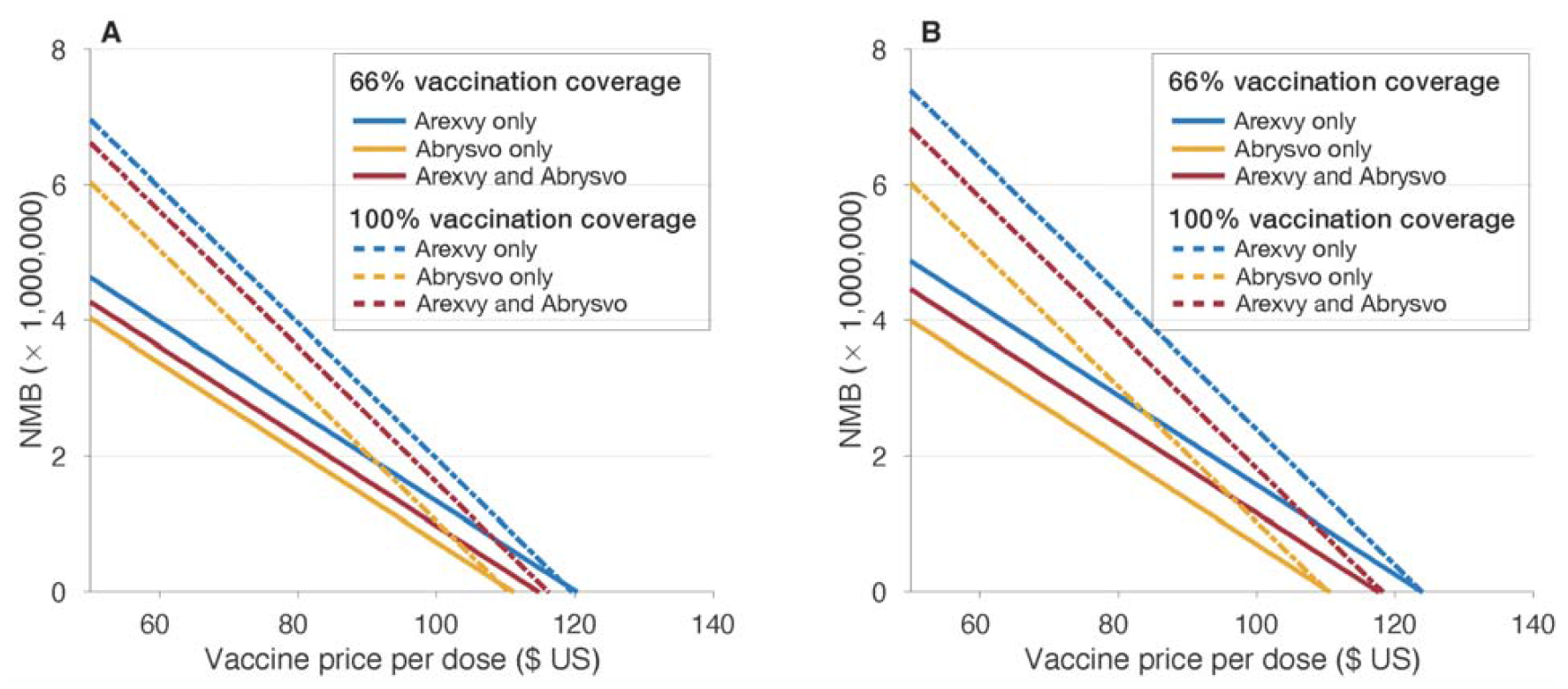
Estimated net monetary benefit (NMB) as a function of price per dose for Arexvy and Abrysvo with different coverage of vaccination. For scenarios using both Arexvy and Abrysvo, each vaccine was assumed to have 50% of the target coverage with the same price per dose.

Under S1, Arexvy at 66% coverage resulted in 46.6 (95% CI: 45.8 to 47.3) QALY gain (**Table 2**). A program with Abrysvo saved 44.3 (95% CI: 43.4 to 44.9) QALYs. Similar estimates were obtained for a combination program using both Arexvy and Abrysvo vaccines (**Table 2**). Increasing vaccination coverage to 100% in S2 resulted in QALY gains of 70.2 (95% CI: 69.5 to 70.9), 66.7 (95% CI: 66.0 to 67.5), and 69.1 (95% CI: 68.4 to 69.8) using Arexvy only, Abrysvo only, and combination of both vaccines, respectively. Sampling from the uncertainty distribution around input parameters, all programs had a 67% probability of being cost-effective at the strategy-specific PPD (**Supplementary Figure A6**).

With constant vaccine efficacy profiles, the maximum PPD for a positive NMB was $123 for Arexvy only, $110 for Abrysvo only, and $117 for a combination of both vaccines in S1 (**Figure 3B**). Under S2, the maximum PPD remained the same as in S1 for each vaccine alone, but changed to $118 for a combination of both vaccines. The estimates of QALY saved were similar to the corresponding scenarios using sigmoidal vaccine efficacy profiles (**Table 2**). The probability of the vaccination program being cost-effective was 67% in all scenarios (**Supplementary Figure A7**).

### Budget impact

With sigmoidal vaccine efficacy profiles, the budget impact to the healthcare system for the first RSV season at the maximum PPD, after discounting for the savings achieved through the reductions of outpatient and inpatient care, was estimated to range from $5.74 to $6.10 billion in S1, and from $8.62 to $9.17 billion in S2 for 74 million older adults in the US (**Table 2**). The budget impact using constant vaccine efficacy profiles were similar and ranged from $5,68 to $6.24 billion in S1, and from $8.61 to $9.43 billion in S2.

### Secondary analysis

Our results for a time-horizon of two RSV seasons (**Supplementary Figure A11**) indicate that the maximum PPD below which vaccination programs are cost-effective at the WTP of $95,000 per QALY depends on the vaccine efficacy. For example, with a sigmoidal decay of vaccine efficacy (**Supplementary Figure A2**), the maximum PPD was $196 for an Arexvy-only program, $186 for an Abrysvo-only program, and $192 when a combination of Arexvy and Abrysvo vaccines were used with 66% vaccination coverage (**Supplementary Table A6**). However, for the constant vaccine efficacy profiles (**Supplementary Figure A3**), the corresponding maximum PPDs for these vaccination programs increased to $250, $233, and $240 (**Supplementary Table A6**). Similar PPDs were estimated with 100% vaccination coverage using sigmoidal and constant vaccine efficacy profiles. At the maximum PPD, all programs were cost-effective with a probability of 67% (**Supplementary Figures A12, A13**). The budget impact of these programs over two years ranged from $8.85 to $17.51 billion for 74 million adults aged 60 years or older depending on the coverage and vaccine efficacy profiles (**Supplementary Table A6**).

## Discussion

In this study, we evaluated the cost-effectiveness of the recently approved prefusion F protein-based RSV vaccines for older adults. We found that vaccination of adults aged 60 years or older could be cost-effective depending on the price as well as the durability of vaccine efficacy. Our results indicate that achieving a 66% vaccination coverage akin to influenza season for older adults would substantially alleviate the burden of RSV-related illness. At the national level, this would require a financial commitment of up to $10.7 billion to cover the costs of purchasing vaccine doses and administration. Beyond the first season, we found that the health benefits of vaccination are sensitive to assumptions about vaccine waning, also affecting the cost-effective PPD and anticipated budget impact over a two-year time horizon.

Although published estimates on efficacy of RSV vaccines are encouraging [6,7], the real-world effectiveness and durability are still unknown. Such estimates are critically important for decision making on effective and cost-effective programs [31], particularly for subpopulations with elevated risk factors. Given the characteristics of the target population with comorbidities, immunocompromised conditions, and immunosenescence, the effectiveness of RSV vaccines in a real-world setting is likely to be lower than those reported in clinical trials. Furthermore, the uncertainty about the benefits of vaccination beyond the first RSV season suggests that the short-term outcomes and the results of cost-effectiveness analysis over a one-year time-horizon would be more appropriate for informing policy decisions on vaccination campaigns.

While the model developed in this study is comprehensive in its structure and accounts for parameter uncertainty at the individual level, there are limitations to consider. Our analysis is based on a discrete-event simulation model, and does not consider the complexity of RSV transmission dynamics. Although no data exist regarding the herd effects of RSV vaccination, it is possible that health benefits could extend beyond just the reduction of disease outcomes by lowering incidence or transmission rates in the population, which could be measured as data become available after the start of mass vaccination. Such data would help parameterize dynamic models of RSV transmission to account for potential indirect benefits of vaccination. Our model does not include costs of potential adverse events post-vaccination, which may affect utility values and could lead to short-term loss of productivity.

In conclusion, our study shows that vaccination against RSV-associated lower respiratory tract disease could be cost-effective and reduce the burden of illness substantially among older adults during the first RSV season. Additional evidence of vaccine effectiveness at the population level would be required to alleviate uncertainty on longer-term health benefits and cost-effectiveness of vaccination beyond a single RSV season.

## Data Availability

All parameter estimautes produced/used in the present work are contained in the manuscript.

https://github.com/affans/rsv-ce-adults

## Authors’ contributions

Seyed Moghadas and Alison Galvani conceived the study; Seyed Moghadas designed the model framework; Carolyn Bawden and Seyed Moghadas collected input parameters; Affan Shoukat developed the computational model and performed simulations; Seyed Moghadas analyzed the simulations data and wrote the first draft of the manuscript; Joanne Langley, Burton Singer, Meagan Fitzpatrick, and Alison Galvani provided insights into the analysis and interpretation of the results; all authors contributed to the writing.

## Reproducibility statement

The computational model is available at: https://github.com/affans/rsv-ce-adults

## Financial support

APG acknowledges support from the NIH grant R01 AI151176, NSF Expeditions grant 1918784, and The Notsew Orm Sands Foundation. SMM acknowledges the support from the NSERC Alliance Grant (ALLRP 576914 - 22). MCF acknowledges support from NIH grant 5 K01 AI141576.

## Potential conflicts of interest

JM Langley’s institution, Dalhousie University, has received funds for clinical trials conducted by the Canadian Center for Vaccinology from GSK, Janssen, Sanofi, Immunovaccine, Inventprise, Merck, Pfizer, VIDO, VBI and Entos. SM Moghadas previously had advisory roles for Janssen Canada and Sanofi for cost-effectiveness of their products. MC Fitzpatrick has received consultation fees from Sanofi Pasteur regarding vaccine products, outside the work reported here.

## Notes

### Competing Interest Statement

JM Langleys institution, Dalhousie University, has received funds for clinical trials conducted by the Canadian Center for Vaccinology from GSK, Janssen, Sanofi, Immunovaccine, Inventprise, Merck, Pfizer, VIDO, VBI and Entos. SM Moghadas previously had advisory roles for Janssen Canada and Sanofi for cost-effectiveness of their products. MC Fitzpatrick has received consultation fees from Sanofi Pasteur regarding vaccine products, outside the work reported here.

### Author Declarations

This study used published estimates and publicly available data sources and thus no ethics approval was required. Consolidated Health Economic Evaluation Reporting Standards (CHEERS) for reporting were followed. The computational model is available at: https://github.com/affans/rsv-ce-adults

