## Supplemental File for "Cost-Effectiveness of Prefusion F Protein-Based Vaccines Against Respiratory Syncytial Virus Disease for Older Adults in the United States"

This supplemental provides additional information for parameterisation of the model, with tables and figures supporting the results described in the main text for cost-effectiveness analysis of vaccination programs over one and two RSV seasons.

**Table A1.** Characteristics of the population age groups.[1,2]

| Age group | Remaining healthy life | Number of comorbidities | | |
|---|---|---|---|---|
| | | 0 | 1–3 | ≥4 |
| 60-64 | 14.8 | 32.0% | 58.2% | 9.8% |
| 65-69 | 11.9 | 26.5% | 62.5% | 11.0% |
| 70-74 | 9.2 | 21.5% | 65.0% | 13.5% |
| 75-79 | 6.8 | 17.0% | 67.5% | 15.5% |
| 80-84 | 4.7 | 14.5% | 67.5% | 18.0% |
| 85+ | 3.1 | 11.5% | 66.5% | 22.0% |

**Table A2.** Model parameters for calculation of productivity loss due to RSV-related outcomes.[3,4]

| Age group | Median income for working individuals (2023) | Labor force participation | Income per day | Working years lost due to death | Loss of productivity due to death | QALY loss due to death |
|---|---|---|---|---|---|---|
| 60-64 | $63,336 | 63.2% | $151.6 | 15 | $778,784 | 9.47 |
| 65-69 | $54,184 | 25.7% | $52.7 | 12 | $555,528 | 7.79 |
| 70-74 | | | | 9 | $434,539 | 5.93 |
| 75-79 | | 8.8% | $18.1 | 7 | $347,709 | 4.49 |
| 80-84 | | | | 5 | $255,591 | 2.97 |
| 85+ | | | | 3 | $157,863 | 1.49 |

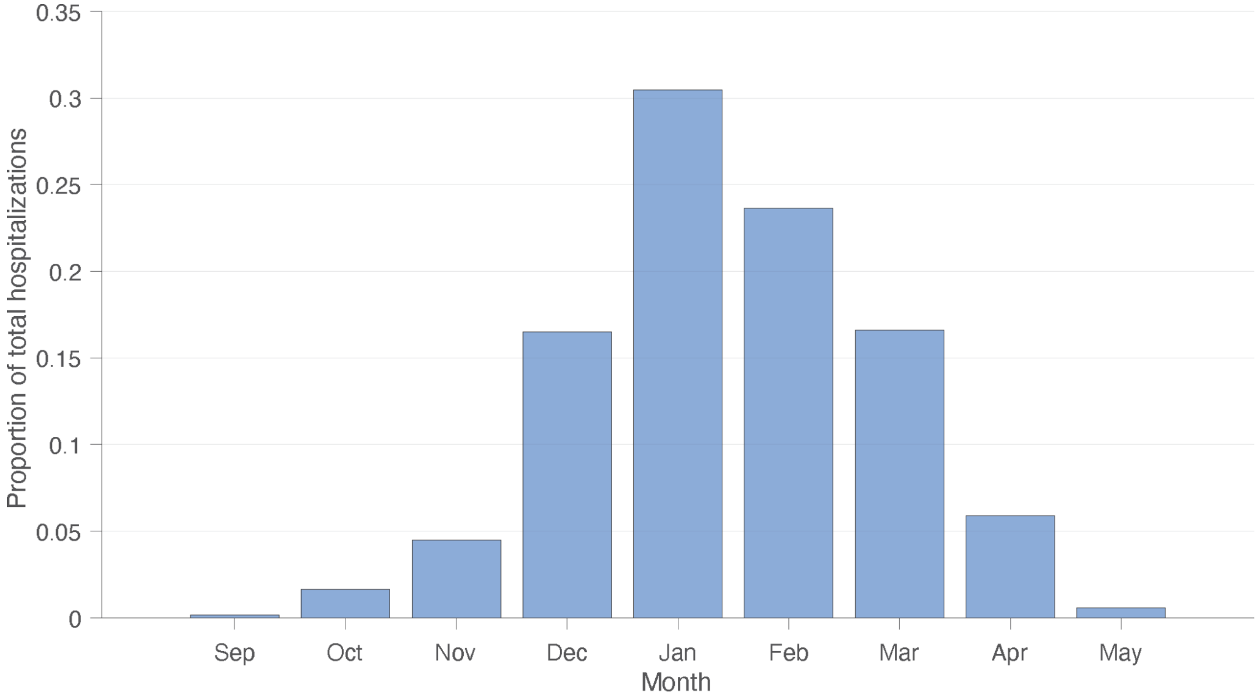


**Figure A1.** Seasonality distribution of RSV-related hospitalizations for adults aged 65 years or older, based on an average of 4 seasons from 2016-17 to 2019-2020 reported in the US.[5] The same distribution was assumed for outpatient care during RSV season.

**Table A3.** Average utility values for population age groups.[2] To calculate the utility values during RSV disease, we applied the utility weights associated with outcomes, corresponding to 0.76 for outpatient care (office or ED visit), 0.35 for hospitalized non-ICU patients, and 0.1 for hospitalized ICU patients.[2,6] The utility weight for non-MA RSV cases during the symptomatic illness was set to 0.88, assuming a 50% lower decrement than MA RSV outpatient care. The duration of effect was assumed to be the same as the duration of outcome.

| Age group | Utility values | | | | |
|---|---|---|---|---|---|
| | Without RSV disease | MA RSV | | | non-MA RSV |
| | | Outpatient | Inpatient non-ICU | Inpatient ICU | |
| 60-64 | 0.77 | 0.5852 | 0.2695 | 0.077 | 0.6776 |
| 65-69 | 0.76 | 0.5776 | 0.2660 | 0.076 | 0.6688 |
| 70-74 | 0.74 | 0.5624 | 0.2590 | 0.074 | 0.6512 |
| 75-79 | 0.70 | 0.532 | 0.2450 | 0.070 | 0.6160 |
| 80-84 | 0.63 | 0.4788 | 0.2205 | 0.063 | 0.5544 |
| 85+ | 0.51 | 0.3876 | 0.1785 | 0.051 | 0.4488 |

**Temporal decline of vaccine efficacy**

To parameterize the model with temporal efficacy of Arexvy and Abrysvo vaccines, we considered a sigmoidal decay function over a 24-month period, given by

$$V_e(t) = \mathrm{VE}_{\mathrm{mean}} - \frac{(\mathrm{VE}_{\mathrm{max}} - \mathrm{VE}_{min})a}{b + e^{-ct}}$$

where $\mathrm{VE}_{\mathrm{mean}}$ is the mean efficacy estimated during the first follow-up period post vaccination, $\mathrm{VE}_{\mathrm{max}}$ and $\mathrm{VE}_{\mathrm{max}}$ are the maximum and minimum efficacy estimates during the entire study period. Assuming that the vaccine efficacy reduced to zero at 24 months after vaccination, we estimated the parameters $a > 0$, $b > 0$, and $c > 0$ (using curve fitting function in Matlab) to derive estimates with the same mean efficacy as estimated in clinical trials.[7,8] Figure A2 illustrates the decline of protection efficacy of Arexvy and Abrysvo over a 24-month period post-dose for different outcomes.

We also considered constant vaccine efficacy profiles with the mean estimates reported in clinical trials over the follow-up period for one and two RSV seasons (Figure A3).[7,8]

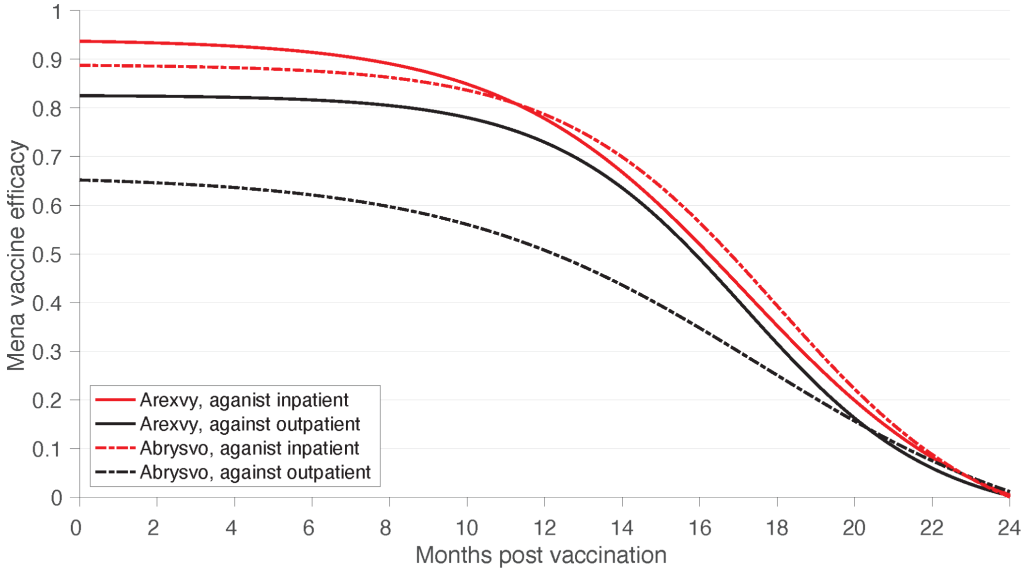


**Figure A2.** Temporal decay of efficacy using sigmoidal fit over 24 months after single dose of Arexvy and Abrysvo vaccines against outpatient (office and ED visits) and inpatient care (hospitalization).

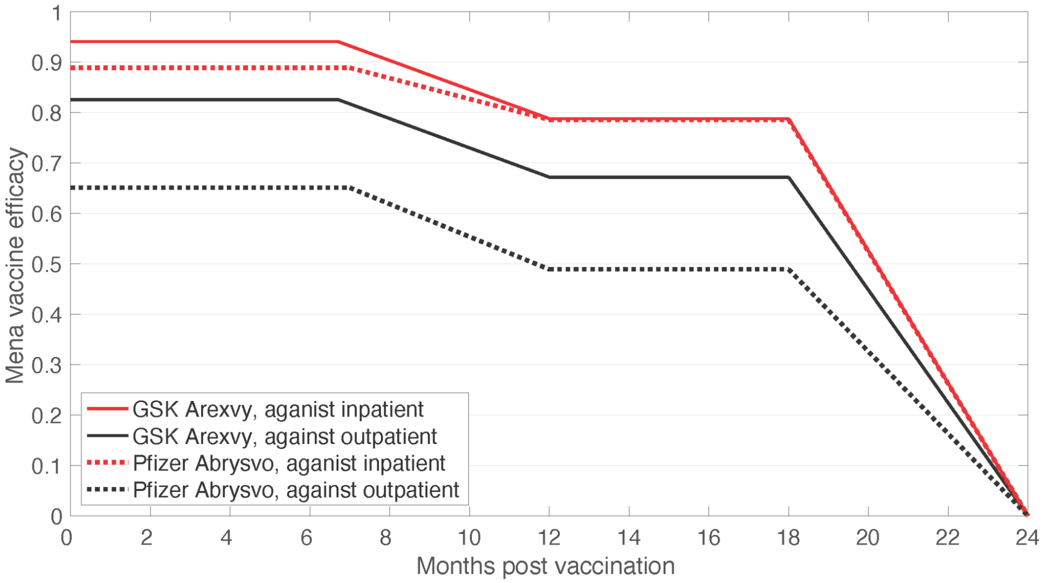


**Figure A3.** Efficacy estimates reported in clinical trials for a single dose of Arexvy and Abrysvo vaccines against outpatient (office and ED visits) and inpatient care (hospitalization) over an 18-month follow-up period. After 18 months, the efficacies were assumed to decline linearly to zero.

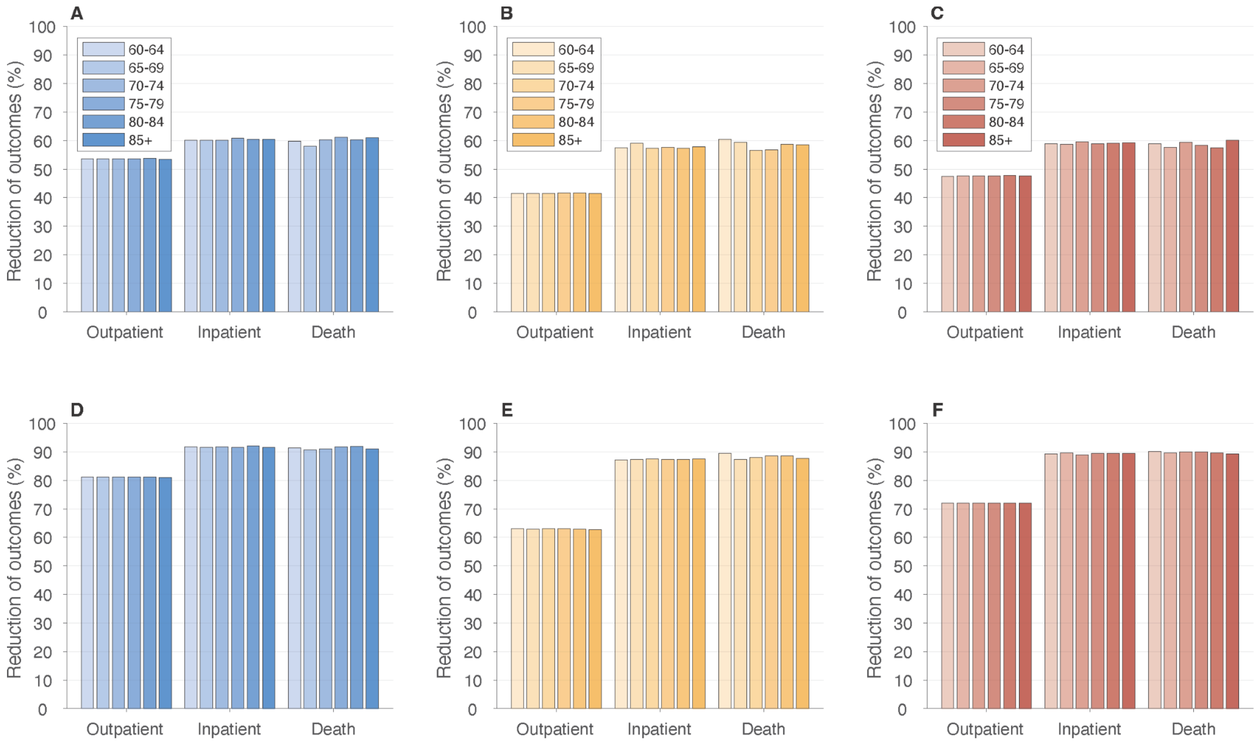


**Figure A4.** Age-specific reductions of outpatient care, hospitalization and deaths achieved in S1 with 66% vaccination coverage (A,B,C) and S2 with 100% vaccination coverage (D,E,F) over a single RSV season. Scenarios correspond to the use of Arexvy vaccine only (A,D); Abrysvo vaccine only (B,E); and a combination of Arexvy and Abrysvo vaccines (C,F), with sigmoidal vaccine efficacy profiles.

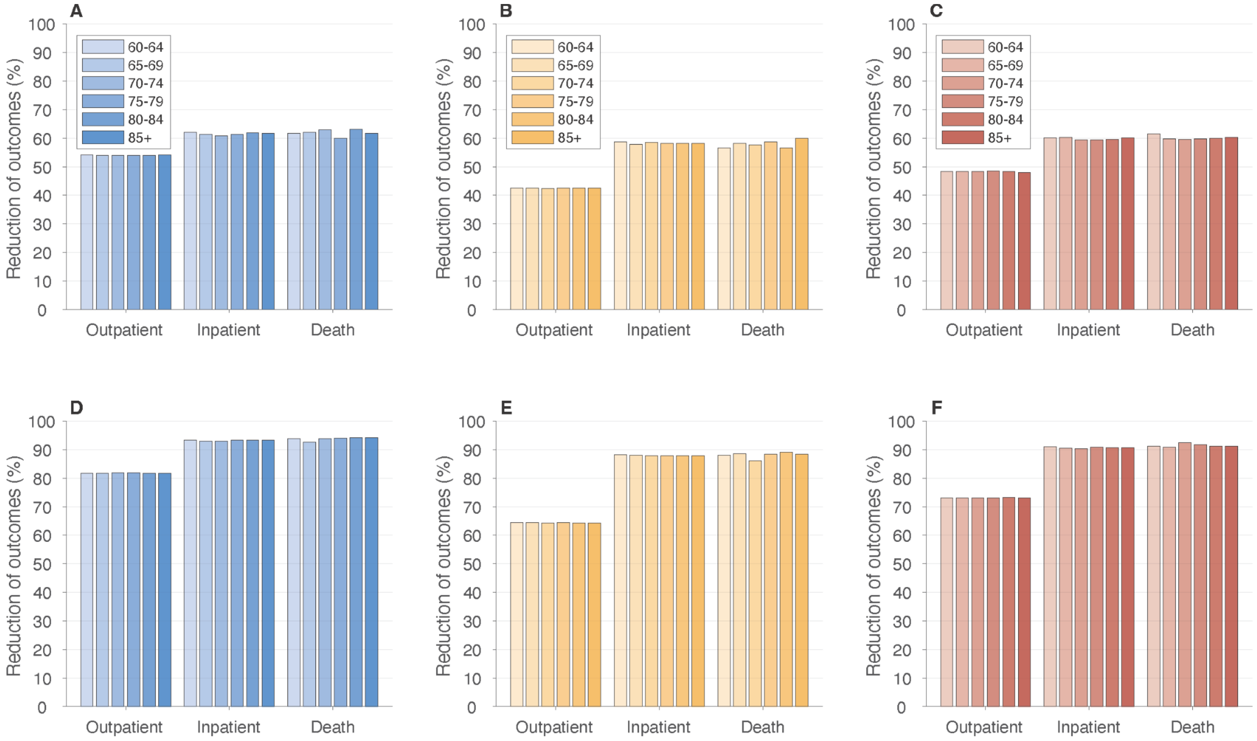


**Figure A5.** Age-specific reductions of outpatient care, hospitalization and deaths achieved in S1 with 66% vaccination coverage (A,B,C) and S2 with 100% vaccination coverage (D,E,F) over a single RSV season. Scenarios correspond to the use of Arexvy vaccine only (A,D); Abrysvo vaccine only (B,E); and a combination of Arexvy and Abrysvo vaccines (C,F), with constant vaccine efficacy profiles.

**Table A4.** Number of vaccine doses needed to avert one outcome over the first RSV season with temporal vaccine efficacy using sigmoidal fit and constant average estimates (**Figures A2 and A3**).

| **Outcome** | **NNV to avert one outcome: mean (95% CI)** | | |
|---|---|---|---|
| | **Arexvy only** | **Abrysvo only** | **Arexvy and Abrysvo** |
| *Sigmoidal vaccine efficacy* | | | |
| Outpatient | 53<br>(52 to 53) | 67<br>(67 to 68) | 60<br>(59 to 60) |
| Hospitalization | 511<br>(507 to 514) | 538<br>(534 to 541) | 520<br>(516 to 524) |
| Death | 7,788<br>(7,667 to 7,917) | 8,108<br>(7,970 to 8,251) | 7,981<br>(7,848 to 8,118) |

| | | | |
|---|---|---|---|
| *Constant vaccine efficacy* | | | |
| Outpatient | 52<br>(52 to 53) | 66<br>(66 to 67) | 58<br>(58 to 59) |
| Hospitalization | 502<br>(498 to 505) | 531<br>(527 to 535) | 516<br>(513 to 520) |
| Death | 7584<br>(7,466 to 7,706) | 8074<br>(7,945 to 8,204) | 7856<br>(7,733 to 7,980) |

### *Cost-effectiveness planes (single RSV season)*

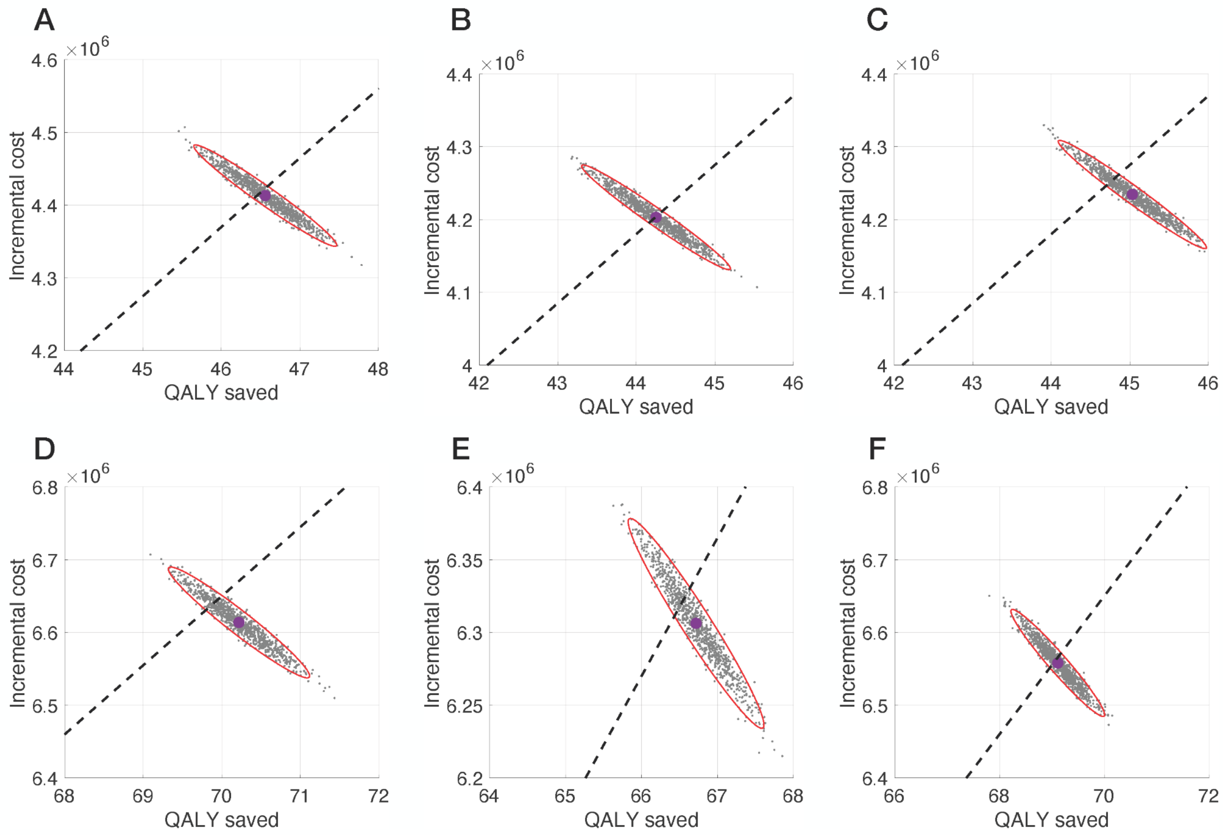


**Figure A6.** Cost-effectiveness planes for vaccination programs with sigmoidal vaccine efficacy profiles for S1 (A,B,C) and S2 (D,E,F). Scenarios correspond to: (A) Arexvy alone with PPD of $120; (B) Abrysvo alone with PPD of $111; a combination of Arexvy and Abrysvo with PPD of $114; (D) Arexvy alone with PPD of $119; (B) Abrysvo alone with PPD of $110; and a combination of Arexvy and Abrysvo with PPD of $116. Black dashed-line corresponds to the WTP threshold of $95,000.

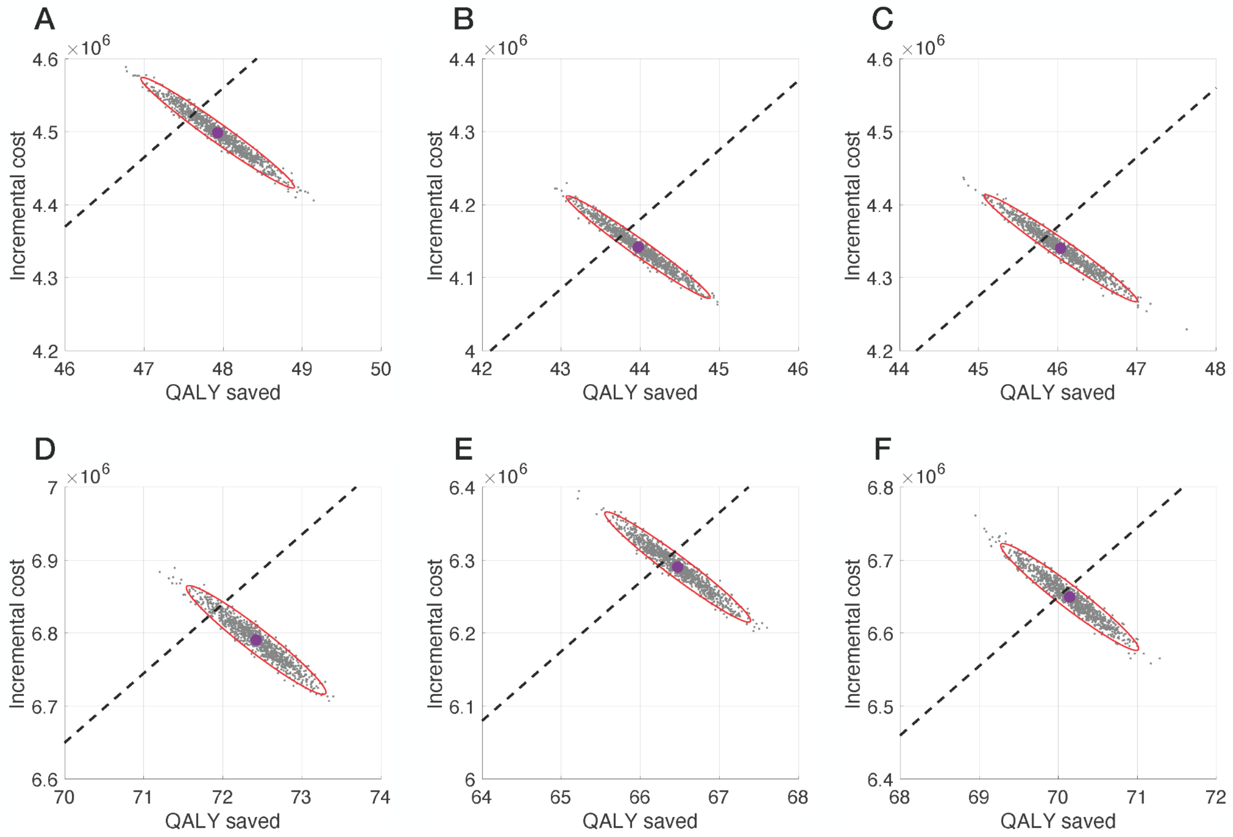


**Figure A7.** Cost-effectiveness planes for vaccination programs with constant vaccine efficacy profiles for S1 (A,B,C) and S2 (D,E,F). Scenarios correspond to: (A) Arexvy alone with PPD of $123; (B) Abrysvo alone with PPD of $110; a combination of Arexvy and Abrysvo with PPD of $117; (D) Arexvy alone with PPD of $123; (B) Abrysvo alone with PPD of $110; and a combination of Arexvy and Abrysvo with PPD of $118. Black dashed-line corresponds to the WTP threshold of $95,000.

### Cost-effectiveness analysis over two RSV seasons

Using sigmoidal vaccine efficacy profile, S1 with 66% vaccination coverage resulted in mean reductions of 42.9%, 32.2%, and 37.5% in outpatient care using Arexvy only, Abrysvo only, and combination of Arexvy and Abrysvo, respectively (**Figure A8A**). The corresponding reductions in hospitalizations were 47.4%, 47.1%, and 47.3%. Similar reductions of 44.1%, 44.2%, and 44.7% in RSV-related deaths were achieved. Increasing vaccination coverage to 100%, S2 resulted in mean reductions of 64.8%, 48.9%, and 56.9% in outpatient care; 71.8%, 71.5%, and 71.7% in hospitalizations; and 66.8%, 67.3%, and 67.3% in deaths using Arexvy only, Abrysvo only, and a combination of Arexvy and Abrysvo, respectively (**Figure A8B**).

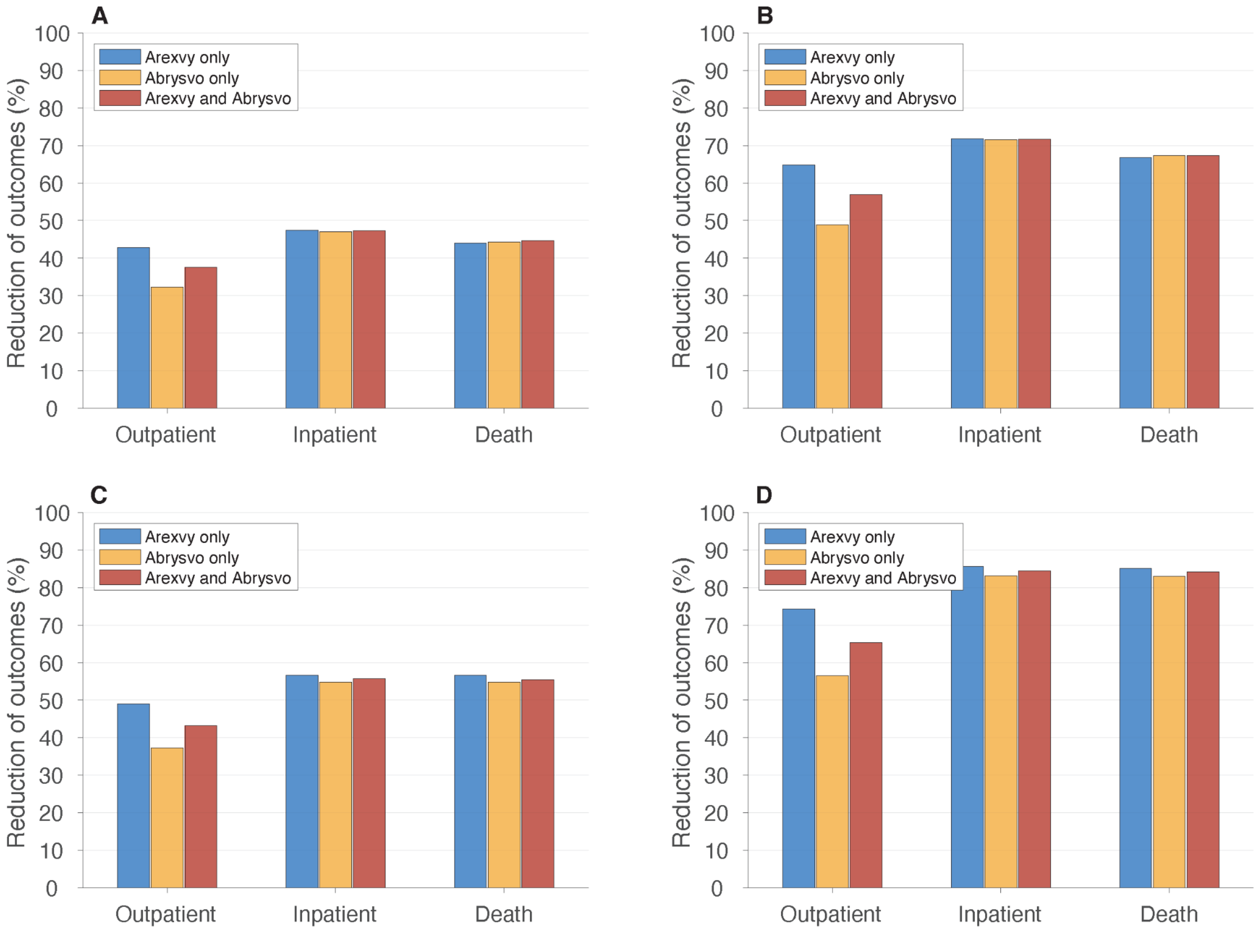


**Figure A8.** Overall reduction of RSV-related outpatient care (office and ED visits), inpatient care (hospitalization),and death among adults 60 years of age or older over two RSV seasons, compared to the scenario without vaccination, with sigmoidal (A,B) and constant (C,D) vaccine efficacy profiles. Vaccination coverage was set to 66% (A,C) and 100% (B,D).

With the constant efficacy profiles, the reduction of outcomes improved. We estimated that S1 with 66% vaccination coverage would reduce outpatient care by 49.1% using Arexvy only, by 37.3% using Abrysvo only, and by 43.2% for a combination of Arexvy and Abrysvo (**Figure A8C**). The corresponding reductions in hospitalizations were 56.7%, 54.8%, and 55.7%. Similar reductions of 56.6%, 54.8%, and 55.4% in RSV-related deaths were achieved. Increasing vaccination coverage to 100%, S2 resulted in mean reductions of 74.3%, 56.6%, and 65.4% in outpatient care; 85.7%, 83.2%, and 84.5% in hospitalizations; and 85.1%, 82.9%, and 84.2% in deaths using Arexvy only, Abrysvo only, and a combination of Arexvy and Abrysvo, respectively (**Figure A8D**).

Similar age-specific reductions of outcomes were achieved for S1 and S2 scenarios using sigmoidal (**Figure A9**) and constant (**Figure A10**) vaccine efficacy profiles.

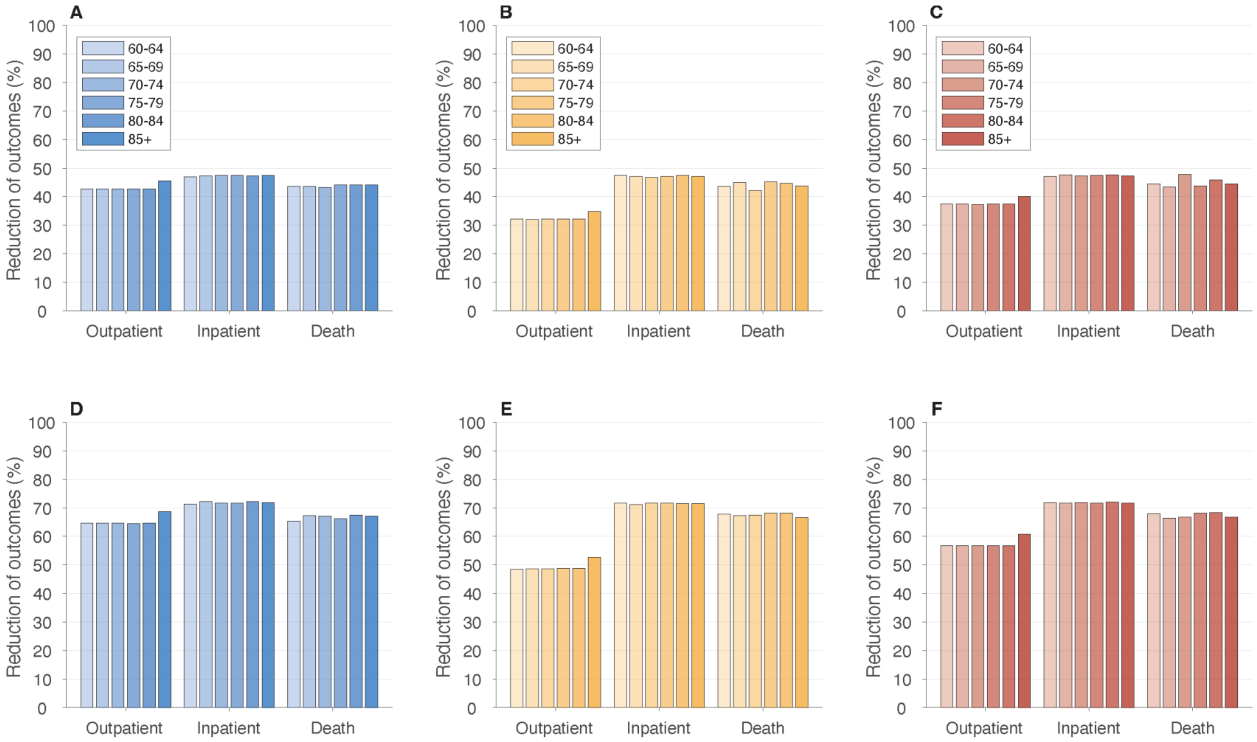


**Figure A9.** Age-specific reductions of outpatient care, hospitalization and deaths achieved in S1 with 66% vaccination coverage (A,B,C) and S2 with 100% vaccination coverage (D,E,F) over a single RSV season. Scenarios correspond to the use of Arexvy vaccine only (A,D); Abrysvo vaccine only (B,E); and a combination of Arexvy and Abrysvo vaccines (C,F), with sigmoidal vaccine efficacy profiles.

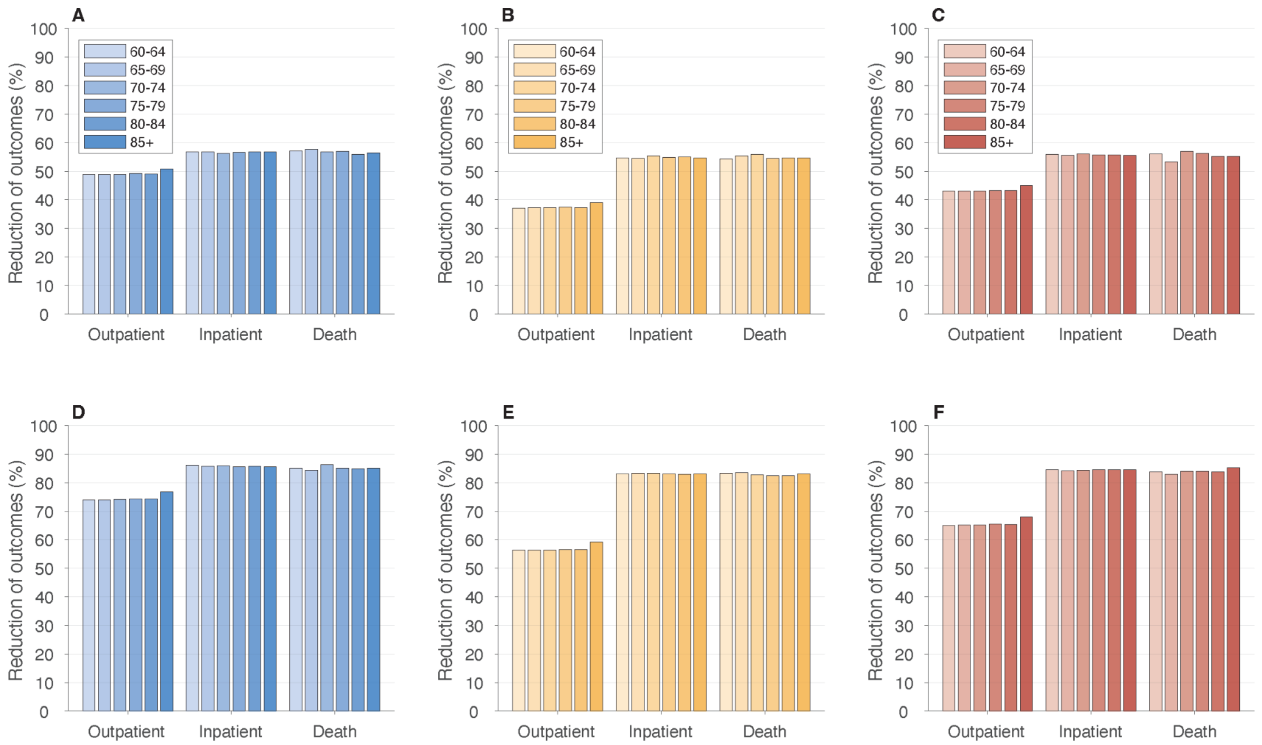


**Figure A10.** Age-specific reductions of outpatient care, hospitalization and deaths achieved in S1 with 66% vaccination coverage (A,B,C) and S2 with 100% vaccination coverage (D,E,F) over a single RSV season. Scenarios correspond to the use of Arexvy vaccine only (A,D); Abrysvo vaccine only (B,E); and a combination of Arexvy and Abrysvo vaccines (C,F), with constant vaccine efficacy profiles.

*Number needed to vaccinate (NNV)*

The mean number of adults in the study population who needed to be vaccinated to avert one outpatient over two RSV seasons with sigmoidal vaccine efficacy profiles ranged from 33 to 38 (**Table A5**). Mean estimated NNV to avert one hospitalization ranged from 324 to 327, and from 5,259 to 5,311 to prevent one death. With the constant vaccine efficacy profiles, NNV to avert one outpatient visit ranged from 29 to 33; to avert one hospitalization ranged from 272 to 281; and to avert one death ranged from 4170 to 4265 (**Table A5**).

**Table A5.** Number of vaccine doses needed to avert one outcome over two RSV seasons with temporal vaccine efficacies using sigmoidal fit and constant average estimates (**Figures A2 and A3**).

| Outcome | NNV to avert one outcome: mean (95% CI) | | |
|---|---|---|---|
| | Arexvy only | Abrysvo only | Arexvy and Abrysvo |

| | | | |
|---|---|---|---|
| *Sigmoidal vaccine efficacy* | | | |
| Outpatient | 33<br>(33 to 33) | 44<br>(43 to 44) | 38<br>(37 to 38) |
| Hospitalization | 324<br>(322 to 325) | 327<br>(325 to 329) | 326<br>(324 to 328) |
| Death | 5,294<br>(5,214 to 5,375) | 5,311<br>(5,233 to 5,390) | 5,259<br>(5,182 to 5,338) |
| *Constant vaccine efficacy* | | | |
| Outpatient | 29<br>(28 to 29) | 38<br>(38 to 38) | 33<br>(33 to 33) |
| Hospitalization | 272<br>(271 to 274) | 281<br>(280 to 283) | 276<br>(275 to 278) |
| Death | 4,170<br>(4,120 to 4,223) | 4,265<br>(4,209 to 4,322) | 4,240<br>(4,186 to 4,297) |

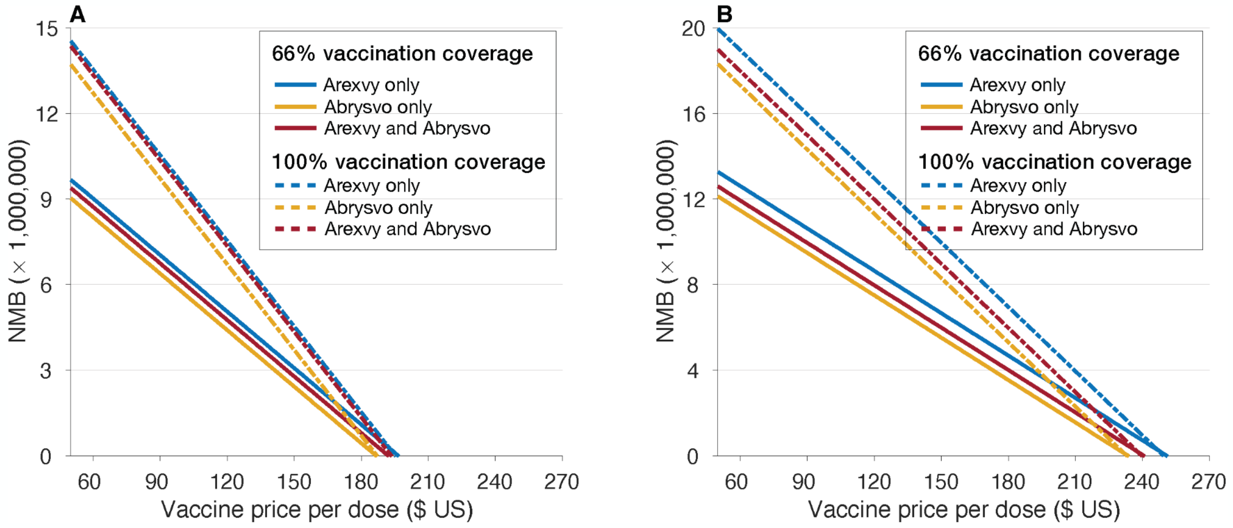


**Figure A11.** Estimated net monetary benefit (NMB) over two RSV seasons as a function of price-per-dose for Arexvy and Abrysvo with different coverage of vaccination. For scenarios using both Arexvy and Abrysvo, each vaccine was assumed to have 50% of the target coverage with the same price-per-dose.

**Table A6.** Model estimates of cost-effectiveness analyses for vaccination programs with Arexvy only, Abrysvo only, and combination of Arexvy and Abrysvo over two RSV seasons in a population of 100,000 adults aged 60 years or older at the WTP of $95,000. All strategies were compared to the baseline with no intervention.

| Scenario | Maximum PPD, $ | Incremental costs, $ (95% CI) | QALYs saved (95% CI) | Budget impact, $ | National budget impact, $ billion |
|---|---|---|---|---|---|
| *S1 with sigmoidal vaccine efficacy* | | | | | |
| Arexvy only | 196 | 6,650,330 (6,569,762 to 6,731,957) | 70.4 (69.3 to 71.5) | 12,521,348 | 9.27 |
| Abrysvo only | 186 | 6,462,916 (6,382,137 to 6,541,894) | 68.5 (67.5 to 69.6) | 11,965,975 | 8.85 |
| Arexvy and Abrysvo | 192 | 6,615,871 (6,545,029 to 6,691,689) | 69.7 (68.7 to 70.7) | 12,318,210 | 9.12 |
| *S2 with sigmoidal vaccine efficacy* | | | | | |
| Arexvy only | 195 | 10,051,020 (9,959,177 to 10,150,313) | 106.0 (104.9 to 107.3) | 18,890,210 | 13.98 |
| Abrysvo only | 187 | 9,886,768 (9,804,336 to 9,974,957) | 104.1 (103.0 to 105.3) | 18,236,324 | 13.49 |
| Arexvy and Abrysvo | 193 | 10,058,133 (9,969,262 to 10,148,406) | 106.4 (105.3 to 107.5) | 18,757,091 | 13.88 |
| *S1 with constant vaccine efficacy* | | | | | |
| Arexvy only | 250 | 8,370,786 (8,283,131 to 8,454,990) | 88.7 (87.6 to 89.9) | 15,667,297 | 11.59 |
| Abrysvo only | 233 | 7,989,830 (7,902,984 to 8,075,577) | 84.5 (83.3 to 85.6) | 14,718,221 | 10.89 |

| | | | | | |
|---|---|---|---|---|---|
| Arexvy and Abrysvo | 240 | 8,127,056<br>(8,043,039 to 8,211,125) | 86.1<br>(84.9 to 87.2) | 15,093,783 | 11.17 |
| *S2 with constant vaccine efficacy* | | | | | |
| Arexvy only | 249 | 12,661,353<br>(12,572,768 to 12,754,502) | 1.338<br>(1.327 to 1.349) | 23,662,539 | 17.51 |
| Abrysvo only | 232 | 12,014,972<br>(11,929,328 to 12,103,220) | 1.275<br>(1.264 to 1.286) | 22,183,068 | 16.42 |
| Arexvy and Abrysvo | 239 | 12,259,374<br>(12,170,858 to 12,338,326) | 1.298<br>(1.288 to 1.310) | 22,775,310 | 16.85 |

*Cost-effectiveness planes (two RSV seasons)*

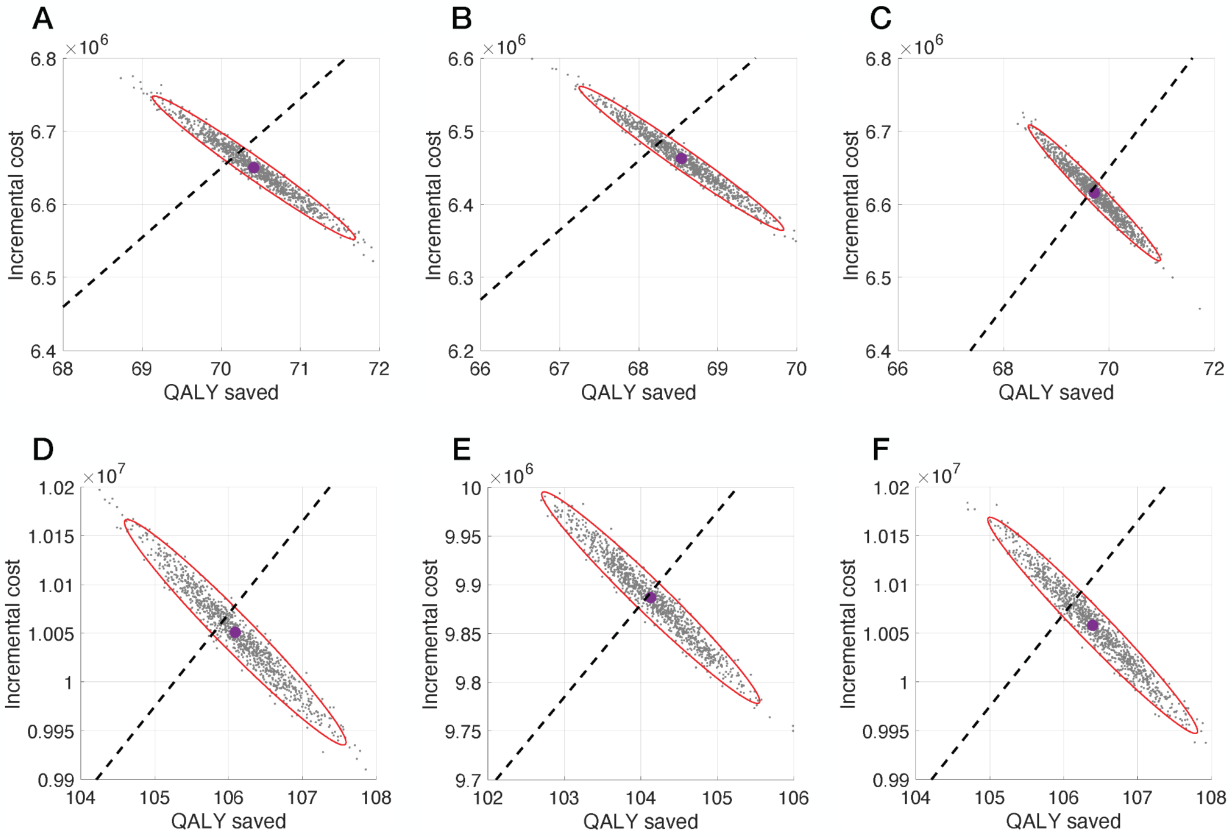


**Figure A12.** Cost-effectiveness planes for vaccination programs over two RSV seasons with sigmoidal vaccine efficacy profiles for S1 (A,B,C) and S2 (D,E,F). Scenarios correspond to: (A) Arexvy alone with PPD of $196; (B) Abrysvo alone with PPD of $186; a combination of Arexvy and Abrysvo with PPD of $192; (D) Arexvy alone with PPD of $195; (B) Abrysvo alone with PPD of $187; and a combination of Arexvy and Abrysvo with PPD of $193. Black dashed-line corresponds to the WTP threshold of $95,000.

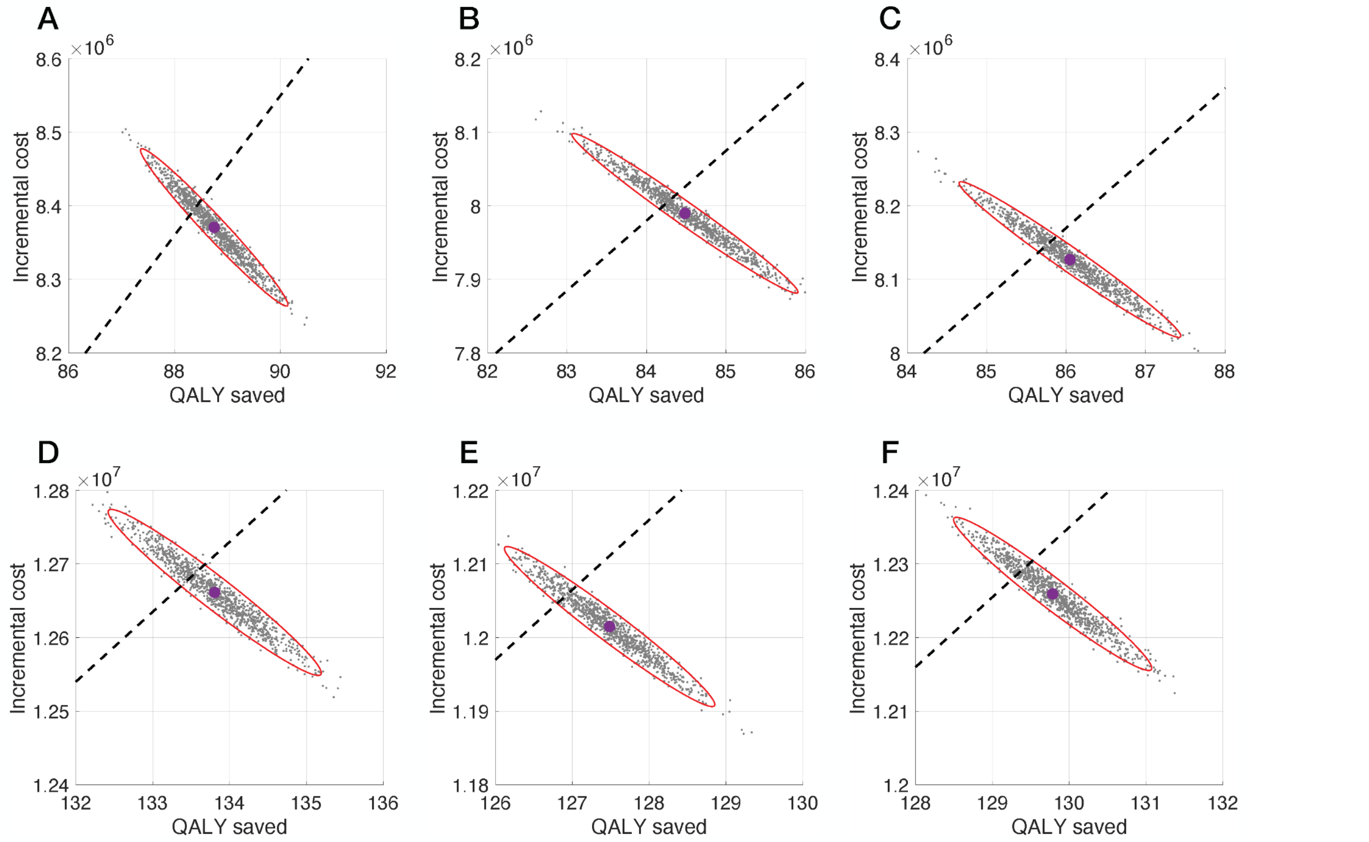


**Figure A13.** Cost-effectiveness planes for vaccination programs over two RSV seasons with constant vaccine efficacy profiles for S1 (A,B,C) and S2 (D,E,F). Scenarios correspond to: (A) Arexvy alone with PPD of $250; (B) Abrysvo alone with PPD of $233; a combination of Arexvy and Abrysvo with PPD of $240; (D) Arexvy alone with PPD of $249; (B) Abrysvo alone with PPD of $232; and a combination of Arexvy and Abrysvo with PPD of $239. Black dashed-line corresponds to the WTP threshold of $95,000.
